# Increasing viral transmission paradoxically reduces progression rates to severe COVID-19 during endemic transition

**DOI:** 10.1101/2022.02.09.22270633

**Authors:** Hyukpyo Hong, Ji Yun Noh, Hyojung Lee, Sunhwa Choi, Boseung Choi, Jae Kyoung Kim, Eui-Cheol Shin

## Abstract

Natural infection with SARS-CoV-2 or vaccination induces virus-specific immunity protecting hosts from infection and severe disease. While the infection-preventing immunity gradually declines, the severity-reducing immunity is relatively well preserved. Here, based on the different longevity of these distinct immunities, we develop a mathematical model to estimate courses of endemic transition of COVID-19. Our analysis demonstrates that high viral transmission unexpectedly reduces the rates of progression to severe COVID-19 during the course of endemic transition despite increased numbers of infection cases. Our study also shows that high viral transmission amongst populations with high vaccination coverages paradoxically accelerates the endemic transition of COVID-19 with reduced numbers of severe cases. These results provide critical insights for driving public health policies in the era of ‘living with COVID-19’.

## Introduction

The coronavirus disease (COVID-19) pandemic is ongoing, resulting in devastating impact on public health, economy, and society. To halt the current pandemic, COVID-19 vaccines have been rapidly developed at an unprecedented pace. These vaccines provide protective immunity against SARS-CoV-2 to prevent infection and limit disease severity, which can also be achieved by natural infection. However, neutralizing antibody (nAb) titers decline after SARS-CoV-2 infection or vaccination in a pattern of initial rapid decay followed by a slower decrease^1,2^, with the half-life of SARS-CoV-2-specific antibodies estimated to be 6∼8 months^1,3^. In addition, SARS-CoV-2 variants exhibit reduced the neutralizing activities of nAb. For example, sera from COVID-19 convalescent patients and vaccine recipients showed reduced neutralizing activities against the Delta (B.1.617.2) and the Omicron (B.1.1.529) variants^4,5^, which became a predominant SARS-CoV-2 strains worldwide. Consequentially, waning humoral immunity to SARS-CoV-2 and the spread of nAb-escaping viral strains (e.g., the Omicron variant) reduce vaccine effectiveness against infection and pose an increasing risk of breakthrough infection over time^6,7^. However, vaccine effectiveness against severe disease is relatively preserved, indicating that different immune components with different half-lives are responsible for preventing infection versus severe disease.

Natural infection or vaccination elicits not only nAbs but also virus-specific CD4^+^ and CD8^+^ memory T cells. nAbs can prevent infection and disease progression by interfering with viral entry to host cells. When hosts are infected, T cells produce effector cytokines and directly eliminate virus-infected cells, leading to rapid control of viral infection and reduction of disease severity. Compared with nAbs, SARS-CoV-2-specific memory T cells are maintained for a relatively long time^8^. Intriguingly, the persistence of memory T-cell responses to SARS-CoV-1 for 17 years has been demonstrated^9^. The long half-life of memory T cells explains the relatively preserved vaccine effectiveness against severe COVID-19^10^.

SARS-CoV-2 is likely to ultimately become endemic and continue circulating among the human population as a common cold virus^11,12^. Indeed, several countries are already considering implementing ‘living with COVID-19’ policies. However, the path to an endemic phase in terms of its duration and public health impact is likely to be highly variable, depending on multiple parameters such as vaccination rates, levels of immunity, transmission rates, and emergence of new variants. Most importantly, being able to control the burden of severe COVID-19 disease, which has the potential to overwhelm health care systems, will be crucial during this transition to an endemic phase.

To enable effective adaptation of public health policies to reduce the overall damage to the community, the future course of the pandemic has been simulated with mathematical models early after the emergence of COVID-19^13–19^. In addition, models incorporating immunity and vaccination were developed^16–18^. In particular, a model demonstrated that infection-induced immunity in children may facilitate endemic transition of COVID-19^19^. However, previous studies did not incorporate the different kinetics of severity-reducing and infection-preventing immunities. Here, we estimate courses of endemic transition of COVID-19 and dynamical changes in progression rates for severe COVID-19 during the transition period, using a mathematical model based on the concept that severity-preventing immunity decay more slowly than infection-preventing immunity.

## Results

We developed a simple mathematical model based on the different kinetics of the two distinct immunities to predict the courses of endemic transition of COVID-19, from the early phase to the steady state (Fig. 1a), including dynamical changes in rates of progression to severe COVID-19 (Fig. 1b, c). Specifically, we extended the Susceptible-Infected-Recovered-Susceptible model, where individuals are separated into five populations: infected individuals with severe disease (*I*_*S*_); infected individuals with mild to moderate disease (*I*_*M*_); individuals susceptible to infection and without immunity (*S*_*H*_), thus a high probability of progression to severe COVID-19 (*h*_*S*_); susceptible individuals with only severity-reducing immunity (*S*_*L*_), thus a low probability of severe COVID-19 progression (*l*_*S*_); and recovered or vaccinated individuals (*R*) with both infection-preventing and severity-reducing immunities (Fig. 1b; see Supplementary Methods for details). Upon SARS-CoV-2 infection with a transmission rate *β, S*_*H*_ and *S*_*L*_ can progress to severe (*I*_*S*_) or mild (*I*_*M*_) COVID-19. Because, the progression rates to severe disease are *h*_*S*_ and *l*_*S*_ for *S*_*H*_ and *S*_*L*_, respectively (Fig. 1c), we refer to 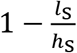 as the efficacy of severity-reducing immunity. A recovery rate is *γ*, and a vaccination rate per day is *v*. Infection-preventing and severity-reducing immunities wane from *R* to *S*_*L*_ and then to *S*_*H*_ at rates 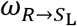 and 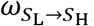, respectively, with 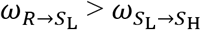 ^12^.

**Fig. 1.**
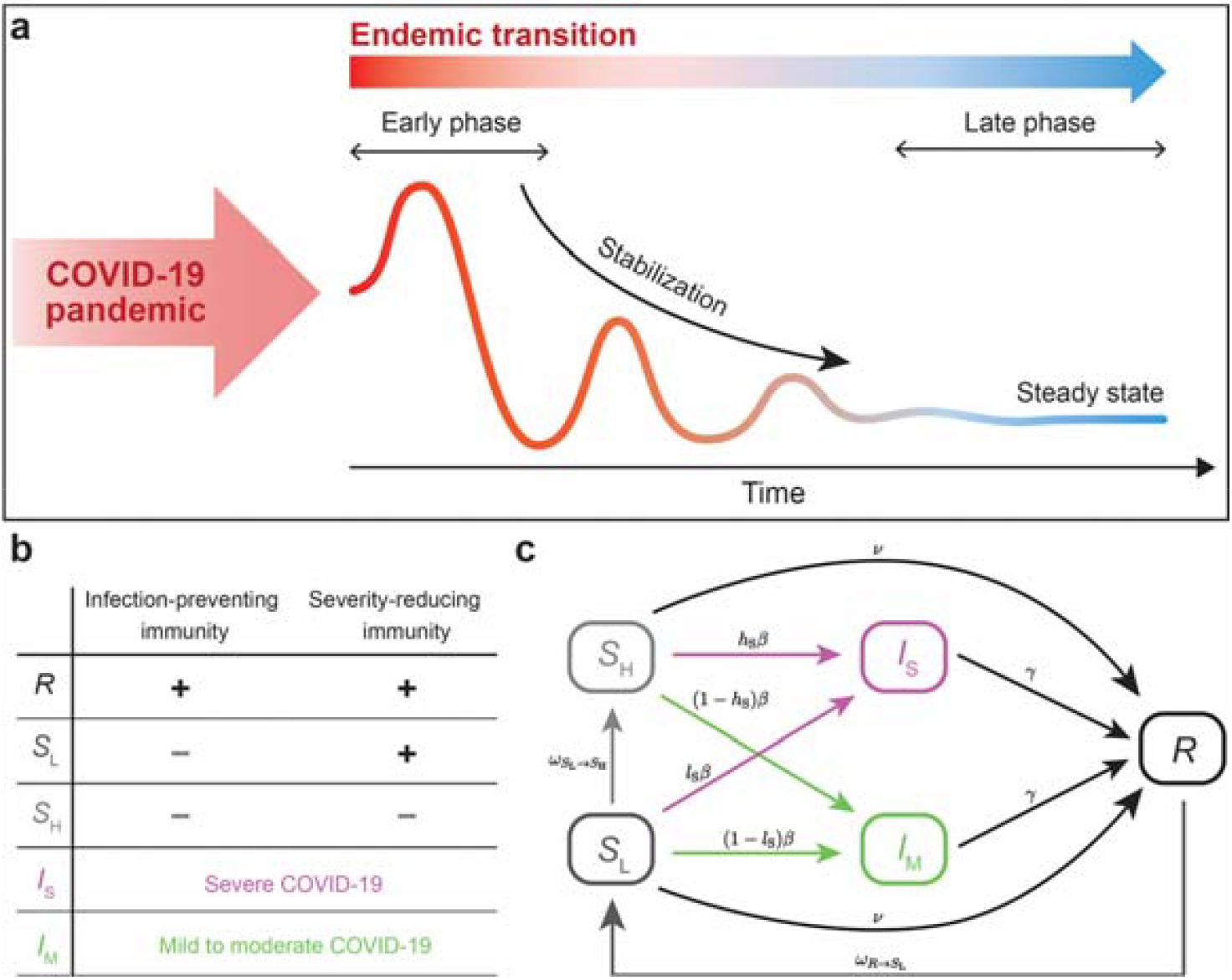
A compartmental model for COVID-19 transmission dynamics incorporating different levels of immunity and disease severity. **a** Schematic illustration for the time-course of endemic transition of the COVID-19 pandemic. **b** The population is divided into five groups: recovered after being infected or vaccinated (*R*); susceptible with a low probability (*S*_*L*_) or a high probability (*S*_*H*_) of experiencing severe disease when they are infected; infected with severe disease (*I*_*S*_), and infected with mild to moderate disease (*I*_*M*_). *R* carry both infection-preventing and severity-reducing immunities, and *S*_*L*_ possesses only severity-reducing immunity. **c** While *S*_*H*_ and *S*_*L*_ can be infected with the same rate *β, S*_*L*_ has a lower rate (*l*_*S*_) of progressing to severe disease (*I*_*S*_) compared to *S*_*H*_ (*h*_*S*_) (i.e., *h*_*S*_ > *l*_*S*_) due to the presence of severity-reducing immunity. *I*_*M*_ and *I*_*S*_ are converted to *R* at a rate *γ*. The infection-preventing and severity-reducing immunities wane at a rate of 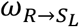 and 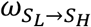, respectively. *S*_*H*_ and *S*_*L*_ can also obtain immunity by vaccination at a rate of *v*.

We first calculated the numbers of daily infections and severe disease at the steady state, i.e., the late phase during endemic transition, according to the level of transmissibility, 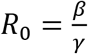 (see Supplementary Methods for details).

When the efficacy of severity-reducing immunity is 95% (*l*_*S*_= 0.05*h*_*S*_), considering vaccine efficacy in preventing severe COVID-19^20^, higher *R*_0_ increases daily infection cases (Fig. 2a, the third panel from the left) as expected. However, higher *R*_0_ decreases the rates of severe disease (Fig. 2b, the third panel from the left) across a wide range of daily vaccination rates. Consequently, the relation between the number of daily severe cases and *R*_0_ is not monotonic (Fig. 2c, the third panel from the left). Specifically, when *R*_0_ is low (*R*_0_ < 1.6), caused for example by strict nonpharmaceutical interventions (NPIs) such as social distancing, the number of daily severe cases decreases with decreasing *R*_0_. However, when *R*_0_ is greater than 1.6, we unexpectedly found that the number of daily severe cases decreases now with increasing *R*_0_. This result is observed when the efficacy of severity-reducing immunity is ≥ 90%, but disappears when it is 80%.

**Fig. 2.**
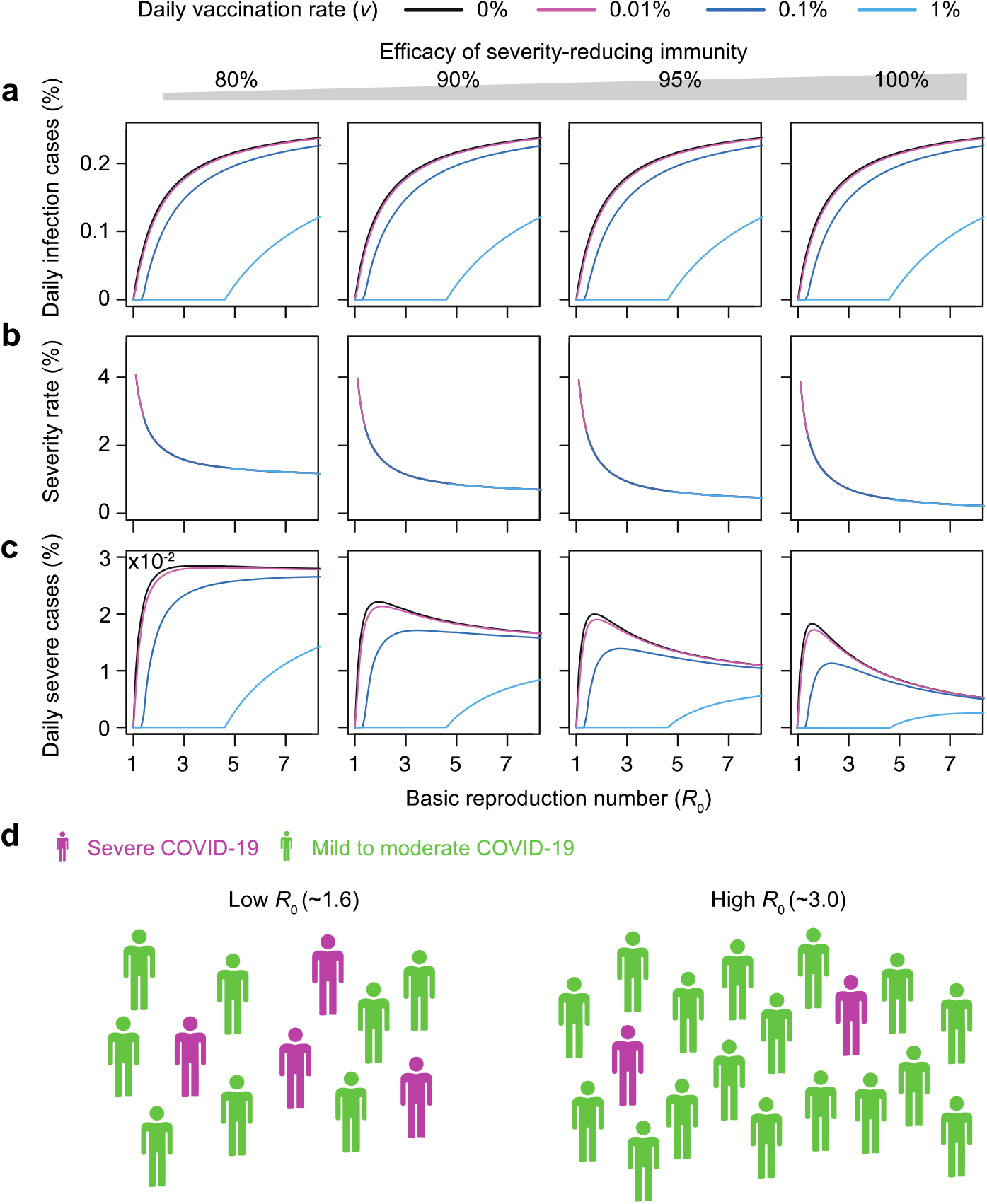
A higher transmission rate can reduce cases of severe disease during the late phase of endemic transition. Steady-state values of the mathematical model over the basic reproduction number 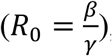, the average number of secondary infections by an infected individual when the whole populations are susceptible, with different daily vaccination rates (*v*) and efficacy of the severity-reducing immunity 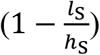. The daily vaccination rates were chosen based on the data of COVID-19 vaccination programs in each country^29^. **a** As transmissibility (*R*_0_) increases, the percentage of daily infections (*γ I*_*S*_ *+ γ I*_*M*_) in the whole populations increases. **b** The percentage of daily infections classified as severe decreases as *R*_*O*_ increases because infection prevents waning of severity-reducing immunity (*S*_*L*_ *→ S*_*H*_). **c** Under strong nonpharmaceutical interventions (NPIs) (*R*_0_<∼1.6), the percentage of severe cases in the whole population increases as *R*_0_ increases. On the other hand, under weak NPIs (*R*_0_>∼1.6), the percentage of severe cases in the whole population decreases as *R*_0_ increases. **d** In summary, higher *R*_0_ increase the daily cases (green + purple) but decreases the severity rate and severe cases (purple). See Supplementary Table 2 for the parameter values.

When *R*_0_ is increased for example by relaxing NPIs or emergence of new variants, the *S*_*L*_ population (i.e., susceptible individuals with severity-reducing immunity) has a high chance of infection but is less likely to have severe disease due to their low rates (*l*_*S*_) for severe disease progression. *S*_*L*_ experiencing re-infection or breakthrough infection is converted to *R* by re-gaining infection-preventing immunity. In summary, high *R*_0_ reduces the proportion of *S*_*H*_ (non-immune population) and maintains individuals within a cycle of *S*_*L*_ ⟷ *R*, leading to a paradoxical decrease in the number of severe cases (*I*_*S*_) at the late phase of endemic transition (Fig. 2d). However, this effect was not observed when a vaccination rate per day is extremely high (Fig. 2, sky blue lines). Similar patterns are demonstrated when we calculate the results in Fig. 2 based on the reproduction number incorporating vaccination (*R*_*v*_; Supplementary Fig. 1) and even when we change values for major parameters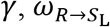, and 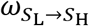 (Supplementary Fig. 2), which have not yet been exactly determined for COVID-19, within a reasonable range (Supplementary Tables 1 and 2).

Although the daily severe cases at the late phase during endemic transition can be reduced by increasing *R*_0_, the number of severe cases will transiently surge during the early phase of endemic transition, which may exceed critical care capacity. Therefore, we investigated transition dynamics during the time-course to an endemic phase under various conditions. When 10% of the population possess infection-preventing immunity, higher *R*_0_ robustly increases the number of daily infection cases during the time-course (Fig. 3a). Although higher *R*_0_ reduces rates for severe disease progression (Fig. 3b), the number of daily severe cases transiently, but sharply increases early during the time-course due to the robust increase in the number of infection cases (Fig. 3c).

**Fig. 3.**
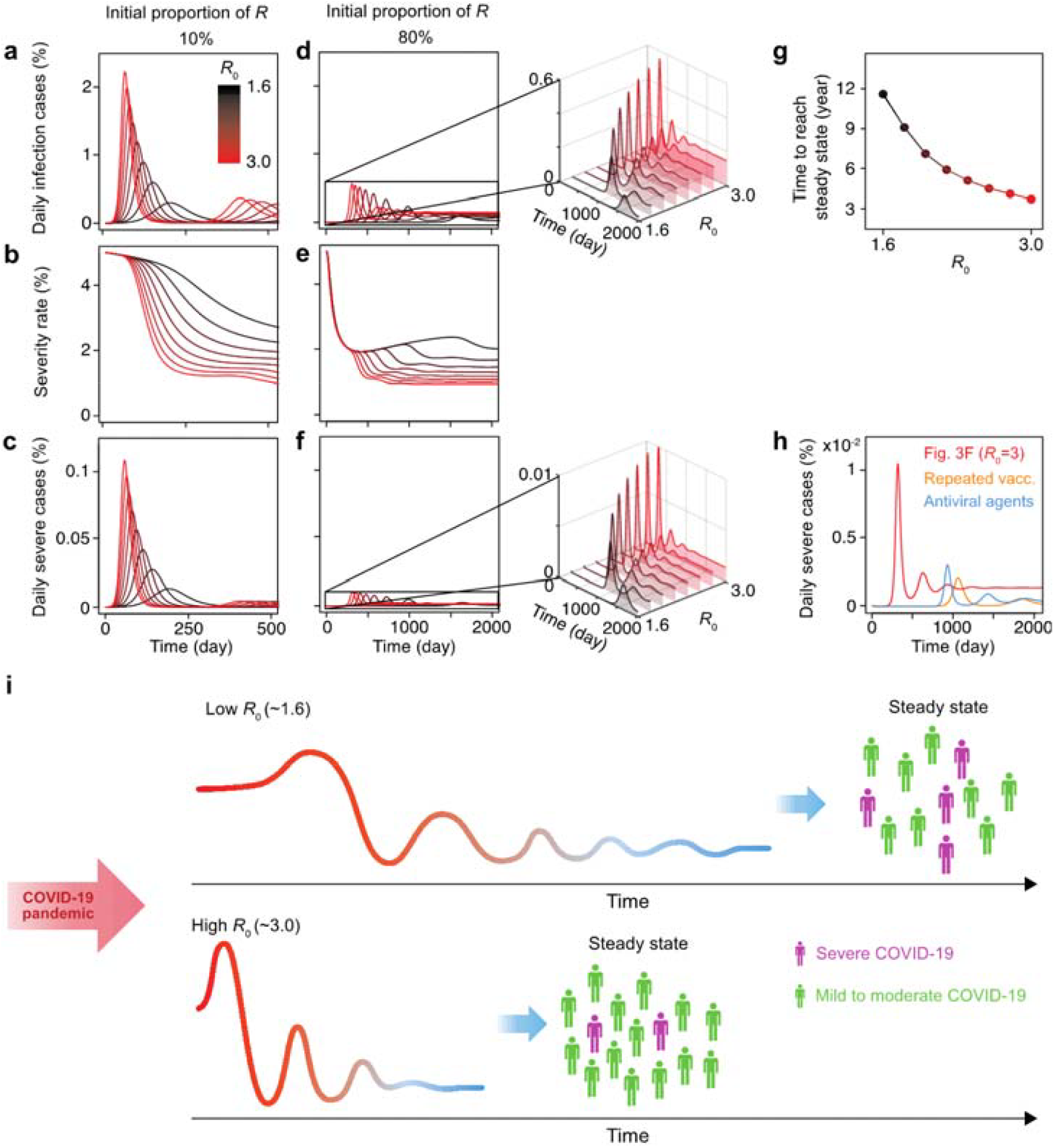
A higher transmission rate can accelerate the transition from the epidemic to the endemic phase without a substantial increase in severe cases. **a** The predicted dynamics of the proportion of daily infection cases in the whole population with varying *R*_0_ from 1.6 to 3.0 for initial infection-preventing immunity of 10% in the population (initial proportion of *R*), acquired by natural infection or vaccination. During the early phase of the endemic transition, higher *R*_0_ increases the daily infection cases. **b, c** Although the percentage of severe cases among all infections becomes lower as *R*_0_ increases (**b**), the surge of percentage of severe cases across the whole population dramatically increases (**c**). (**d-f**) Predicted transition dynamics are shown for higher initial infection-preventing immunity of 80% (initial proportion of *R*). The surge of severe cases is greatly reduced compared with when the initial immunity is 10% (**f**), because both infection cases (**d**) and rate of severe cases (**e**) decrease. Furthermore, higher *R*_0_ accelerates the stabilization of the number of daily cases (**d**) and severe cases (**f**). **g** Time to reach the stage when the number of severe cases fluctuates between 70% and 130% of the steady-state value. **h** The percentage of severe cases when two interventions are implemented. The red line is recalled from (**f**) when *R*_0_=3.0, the orange line is dynamics from reduced waning rates (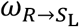 and 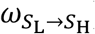), and the blue line is dynamics from increased recovery rate (*γ*) and decreased probability of experiencing severe disease (*h*_*S*_ and *l*_*S*_). See Supplementary Table 2 for the parameter values. **i** A graphical summary of the results. Compared with the low *R*_0_, the high *R*_0_ leads to an earlier endemic transition and a reduced severity rate and the number of severe cases.

The spike of severe cases is dramatically attenuated when a high proportion of the population (e.g., 80%) possess infection-preventing immunity, by natural infection or vaccination (Fig. 3d-f). Moreover, with higher *R*_0_, the curves of severe cases are stabilized more quickly to the steady state without fluctuation (Fig. 3f). Times to reach the steady state are estimated to be 12 and 4 years with *R*_0_ *=* 1.6 and 3.0, respectively (Fig. 3g). When the proportion of the population with infection-preventing immunity is 50%, similar patterns are observed while peaks of infection and severe cases occur earlier and are higher compared to 80% immunity (Supplementary Fig. 3). This demonstrates that permissive viral spread under high vaccination coverages accelerates the endemic transition with subsequent controllable COVID-19 disease burden. However, sharp peaks of severe cases in the early phase could still threaten overwhelming healthcare systems.

To circumvent this, we evaluated the effects of repeated vaccinations (i.e., boosters) and antiviral agents on the curves of severe cases. Repeated vaccinations, which increase the longevity of both infection-preventing and severity-reducing immunities (i.e., decrease in 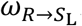 and 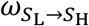)^21,22^, reduced the peak of severe cases (Fig. 3h, orange line). Indeed, an additional third dose of the mRNA vaccine showed a 92% effectiveness in preventing severe COVID-19 disease compared with two-dose vaccination^23^. Novel antiviral agents, which will increase the recovery rate *γ* and decrease progression to severe disease, *h*_*S*_ and *l*_*S*_, also reduced the peak of severe cases (Fig. 3h, blue line). In fact, molnupiravir, a newly developed antiviral agent, has been shown to reduce the risk of hospitalization or death by 30% in patients with mild-to-moderate COVID-19^24^.

## Discussion

As the COVID-19 pandemic is ongoing, predicting the future course of the pandemic is needed to enable effective adaptation of public health policies to reduce the overall damage to the community. The concept of an endemic transition of SARS-CoV-2 has been proposed^11,12,19^, however the possible impact of severe COVID-19 cases during this transition has not been estimated. This is crucial for driving appropriate public health policies and ensuring that healthcare systems can subsequently withstand the disease burden.

For this, we developed a simple model focusing on two heterogeneous features: immunity and clinical severity. This allowed us to forecast courses of endemic transition of COVID-19 based on the concept that severity-preventing immunity decays more slowly than infection-preventing immunity. In particular, increasing viral spread, for example by relaxing NPIs, under high vaccination coverages paradoxically reduces progression rates to severe COVID-19 and stabilizes the development of severe cases during the transition to an endemic phase, with reduced numbers of severe cases.

Emergence of new SARS-CoV-2 variants can also change the course of the endemic transition and the number of severe cases. Natural selection of mutant viruses occurs under the pressure of increasing viral fitness or escaping from immunity. New variants with higher fitness that more efficiently enter host cells or replicate can change the course of endemic transition by increasing *R*_0_. In the case of immune-evading variants, both 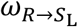 and 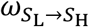 can be increased in theory. Given that nAbs prevent infection by interfering with viral entry and are easily evaded by variants, the emergence of variants can reduce infection-preventing immunity and increase 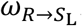. However, variants rarely escape SARS-CoV-2-specific memory T cell responses that can prevent severe disease because T-cell epitopes are scattered across the viral proteome, suggesting that the emergence of variants minimally changes severity-reducing immunity or 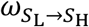.

Recently, the Omicron variant (B.1.1.529), a new variant of concern harbouring the high number of mutations in the spike protein, has emerged^25^. The Omicron variant was estimated to have higher reproduction number than the Delta variant^26^. It was also experimentally demonstrated that Omicron spike-pseudovirus exhibits greater efficiency of target cell entry than other SARS-CoV-2 pseudoviruses^27^. Moreover, the Omicron variant has been shown to reduce the neutralizing activities of nAbs elicited by COVID-19 vaccination or infection with other SARS-CoV-2 strains^5,27^. However, the Omicron variant is known to result in less severe infection than other SARS-CoV-2 strains. Low pathogenicity of the Omicron variant is explained by preferential infection of the upper airway rather than the lungs^28^. Such distinct characteristics of the Omicron variant can be flexibly incorporated into our model. Importantly, spread of the Omicron variant with high transmissibility is likely to facilitate the endemic transition of COVID-19 according to our model prediction.

In the current study, to predict the qualitative trends of COVID-19 endemic transition, we developed a simple model focusing on two heterogeneous features: immunity and clinical severity, while simplifying other population heterogeneities such as age, underlying disease, and cross-reactive immunity elicited by other coronaviruses as previous work^17^. These heterogeneities need to be incorporated for precise quantitative prediction. Our model allowed us to forecast courses of endemic transition of COVID-19 based on the concept that severity-reducing immunity decays more slowly than infection-preventing immunity. We demonstrate that increasing viral spread, for example by relaxing NPIs or emergence of new variants, under high vaccination coverages paradoxically reduces progression rates to severe COVID-19 and stabilizes the development of severe cases during the endemic transition, with reduced numbers of severe cases (Fig. 3i). While our prediction needs to be interpreted appropriately depending on each country, it provides important insights for establishing or adjusting public health policies in the era of ‘living with COVID-19’.

## Methods

### Model parameters

We have used a plausible range for model parameters from the published literature (Supplementary Table 1) rather than a single parameter set to get robust model prediction against parameter perturbation.

### Reproduction numbers and steady state formulae derivation

We derived formula for the basic reproduction number *R*_0_, the average number of secondary infections by an infected individual when the whole populations are susceptible (see Supplementary Methods for details). A reproduction number incorporating vaccination was also derived similarly. Furthermore, we derived the steady-state values of the variables in the model for a given set of parameters (see Supplementary Methods for details). It allowed us to obtain the late-phase status without performing numerical simulation of the model.

### Data availability

Code and data used to perform the analysis and generate the figures in this paper are available on GitHub (https://github.com/Mathbiomed/EndemicSIRS).

## Data Availability

All data produced in the present work are contained in the manuscript.

https://github.com/Mathbiomed/EndemicSIRS

## Acknowledgments

The authors thank Life Science Editors for editing support. This work was supported by the Institute for Basic Science (IBS-R801-D2 to E.-C.S. and IBS-R029-C3 to J.K.K.), Korea Health Technology R&D Project through the Korea Health Industry Development Institute, funded by the Ministry of Health & Welfare, Republic of Korea HI20C0452 (J.Y.N.), National Research Foundation of Korea (NRF-2021R1A2C1095639 to S.C., NRF-2020R1F1A1A01066082 to B.C., and 2019H1A2A1075303 to H.H.)

## Author contributions

Conceptualization: H.H., J.Y.N., J.K.K., E.-C.S.

Methodology: H.H., J.Y.N., H.L., S.C., B.C., J.K.K., E.-C.S.

Investigation: H.H., J.Y.N., H.L., S.C., B.C., J.K.K., E.-C.S.

Visualization: H.H., J.Y.N., J.K.K., E.-C.S.

Funding acquisition: H.H., J.Y.N., S.C., B.C., J.K.K., E.-C.S.

Supervision: J.K.K., E.-C.S.

Writing – original draft: H.H., J.Y.N., J.K.K., E.-C.S.

Writing – review & editing: H.H., J.Y.N., H.L., S.C., B.C., J.K.K., E.-C.S.

## Competing interests

Authors declare that they have no competing interests.

## Notes

### Competing Interest Statement

The authors have declared no competing interest.

## References

1 Cohen, K. W. et al. Longitudinal analysis shows durable and broad immune memory after SARS-CoV-2 infection with persisting antibody responses and memory B and T cells. Cell Rep. Med. 2, 100354, doi:10.1016/j.xcrm.2021.100354 (2021).

2 Levin, E. G. et al. Waning Immune Humoral Response to BNT162b2 Covid-19 Vaccine over 6 Months. N. Engl. J. Med., doi:10.1056/NEJMoa2114583 (2021).

3 Doria-Rose, N. et al. Antibody Persistence through 6 Months after the Second Dose of mRNA-1273 Vaccine for Covid-19. N. Engl. J. Med. 384, 2259–2261, doi:10.1056/NEJMc2103916 (2021).

4 Liu, C. et al. Reduced neutralization of SARS-CoV-2 B.1.617 by vaccine and convalescent serum. Cell 184, 4220–4236.e4213, doi:10.1016/j.cell.2021.06.020 (2021).

5 Liu, L. et al. Striking Antibody Evasion Manifested by the Omicron Variant of SARS-CoV-2. Nature, doi:10.1038/s41586-021-04388-0 (2021).

6 Chemaitelly, H. et al. Waning of BNT162b2 Vaccine Protection against SARS-CoV-2 Infection in Qatar. N. Engl. J. Med., doi:10.1056/NEJMoa2114114 (2021).

7 Mizrahi, B. et al. Correlation of SARS-CoV-2-breakthrough infections to time-from-vaccine. Nat. Commun. 12, 6379, doi:10.1038/s41467-021-26672-3 (2021).

8 Jung, J. H. et al. SARS-CoV-2-specific T cell memory is sustained in COVID-19 convalescent patients for 10 months with successful development of stem cell-like memory T cells. Nat. Commun. 12, 4043, doi:10.1038/s41467-021-24377-1 (2021).

9 Le Bert, N. et al. SARS-CoV-2-specific T cell immunity in cases of COVID-19 and SARS, and uninfected controls. Nature 584, 457–462, doi:10.1038/s41586-020-2550-z (2020).

10 Noh, J. Y., Jeong, H. W., Kim, J. H. & Shin, E. C. T cell-oriented strategies for controlling the COVID-19 pandemic. Nat. Rev. Immunol. 21, 687–688, doi:10.1038/s41577-021-00625-9 (2021).

11 Veldhoen, M. & Simas, J. P. Endemic SARS-CoV-2 will maintain post-pandemic immunity. Nat. Rev. Immunol. 21, 131–132, doi:10.1038/s41577-020-00493-9 (2021).

12 Antia, R. & Halloran, M. E. Transition to endemicity: Understanding COVID-19. Immunity 54, 2172–2176, doi:10.1016/j.immuni.2021.09.019 (2021).

13 Lovell-Read, F. A., Funk, S., Obolski, U., Donnelly, C. A. & Thompson, R. N. Interventions targeting non-symptomatic cases can be important to prevent local outbreaks: SARS-CoV-2 as a case study. J. R. Soc. Interf. 18, 20201014, doi:10.1098/rsif.2020.1014 (2021).

14 Sahai, S. Y., Gurukar, S., KhudaBukhsh, W. R., Parthasarathy, S. & Rempala, G. A. A machine learning model for nowcasting epidemic incidence. Math. Biosci., 108677, doi:10.1016/j.mbs.2021.108677 (2021).

15 Kucharski, A. J. et al. Early dynamics of transmission and control of COVID-19: a mathematical modelling study. Lancet Infect. Dis. 20, 553–558, doi:10.1016/S1473-3099(20)30144-4 (2020).

16 Kissler, S. M., Tedijanto, C., Goldstein, E., Grad, Y. H. & Lipsitch, M. Projecting the transmission dynamics of SARS-CoV-2 through the postpandemic period. Science 368, 860–868, doi:10.1126/science.abb5793 (2020).

17 Saad-Roy, C. M. et al. Epidemiological and evolutionary considerations of SARS-CoV-2 vaccine dosing regimes. Science 372, 363–370, doi:10.1126/science.abg8663 (2021).

18 Saad-Roy, C. M. et al. Immune life history, vaccination, and the dynamics of SARS-CoV-2 over the next 5 years. Science 370, 811–818, doi:10.1126/science.abd7343 (2020).

19 Lavine, J. S., Bjornstad, O. N. & Antia, R. Immunological characteristics govern the transition of COVID-19 to endemicity. Science 371, 741–745, doi:10.1126/science.abe6522 (2021).

20 Thomas, S. J. et al. Safety and Efficacy of the BNT162b2 mRNA Covid-19 Vaccine through 6 Months. N. Engl. J. Med. 385, 1761–1773, doi:10.1056/NEJMoa2110345 (2021).

21 Falsey, A. R. et al. SARS-CoV-2 Neutralization with BNT162b2 Vaccine Dose 3. N. Engl. J. Med. 385, 1627–1629, doi:10.1056/NEJMc2113468 (2021).

22 Bertrand, D. et al. Antibody and T-cell response to a third dose of SARS-CoV-2 mRNA BNT162b2 vaccine in kidney transplant recipients. Kidney Int. 100, 1337–1340, doi:10.1016/j.kint.2021.09.014 (2021).

23 Barda, N. et al. Effectiveness of a third dose of the BNT162b2 mRNA COVID-19 vaccine for preventing severe outcomes in Israel: an observational study. Lancet, doi:10.1016/s0140-6736(21)02249-2 (2021).

24 Jayk Bernal, A. et al. Molnupiravir for Oral Treatment of Covid-19 in Nonhospitalized Patients. N. Engl. J. Med., doi:10.1056/NEJMoa2116044 (2021).

25 Kumar, S., Thambiraja, T. S., Karuppanan, K. & Subramaniam, G. Omicron and Delta variant of SARS-CoV-2: A comparative computational study of spike protein. J. Med. Virol., doi:10.1002/jmv.27526 (2021).

26 Nishiura, H. et al. Relative Reproduction Number of SARS-CoV-2 Omicron (B.1.1.529) Compared with Delta Variant in South Africa. J. Clin. Med. 11, doi:10.3390/jcm11010030 (2021).

27 Garcia-Beltran, W. F. et al. mRNA-based COVID-19 vaccine boosters induce neutralizing immunity against SARS-CoV-2 Omicron variant. Cell, doi:10.1016/j.cell.2021.12.033 (2022).

28 Kozlov, M. Omicron’s feeble attack on the lungs could make it less dangerous. Nature 601, 177, doi:10.1038/d41586-022-00007-8 (2022).

29 Ritchie, H. et al. Coronavirus Pandemic (COVID-19). (2020). https://ourworldindata.org/coronavirus (accessed Jan 18, 2022)

