## Supplementary Methods and Figures for "Increasing viral transmission paradoxically reduces progression rates to severe COVID-19 during endemic transition"

#### The description of the mathematical model (Fig. 1c)

The evolution of epidemics for SARS-Cov2 can be described by the following system of ordinary differential equations (see Supplementary Table 1 for the description of parameters):

$$\frac{dS_{\mathrm{H}}}{dt} = -\beta\frac{(I_{\mathrm{S}}+I_{\mathrm{M}})}{N}S_{\mathrm{H}} - \nu S_{\mathrm{H}} + \omega_{S_{\mathrm{L}}\to S_{\mathrm{H}}}S_{\mathrm{L}},$$
$$\frac{dS_{\mathrm{L}}}{dt} = -\beta\frac{(I_{\mathrm{S}}+I_{\mathrm{M}})}{N}S_{\mathrm{L}} - \nu S_{\mathrm{L}} - \omega_{S_{\mathrm{L}}\to S_{\mathrm{H}}}S_{\mathrm{L}} + \omega_{R\to S_{\mathrm{L}}}R,$$
$$\frac{dI_{\mathrm{S}}}{dt} = \beta\frac{(I_{\mathrm{S}}+I_{M})}{N}(h_{\mathrm{S}}S_{\mathrm{H}} + l_{\mathrm{S}}S_{\mathrm{L}}) - \gamma I_{\mathrm{S}},$$
$$\frac{dI_{\mathrm{M}}}{dt} = \beta\frac{(I_{\mathrm{S}}+I_{M})}{N}((1-h_{\mathrm{S}})S_{\mathrm{H}} + (1-l_{\mathrm{S}})S_{\mathrm{L}}) - \gamma I_{\mathrm{M}},$$
$$\frac{dR}{dt} = \gamma(I_{\mathrm{S}}+I_{M}) + \nu(S_{\mathrm{H}}+S_{\mathrm{L}}) - \omega_{R\to S_{\mathrm{L}}}R,$$

where $S_{\mathrm{H}}$, $S_{\mathrm{L}}$, $I_{\mathrm{S}}$, $I_{\mathrm{M}}$, and $R$ represent susceptible individuals without immunity, susceptible individuals with severity-reducing immunity, infected individuals with severe disease, infected individuals with mild to moderate disease, and recovered individuals with infection-preventing immunity, respectively. In the model, those in the susceptible population (i.e., $S_{\mathrm{H}}$ and $S_{\mathrm{L}}$) are infected via contact with infectious individuals (i.e., $I_{\mathrm{S}}$ and $I_{\mathrm{M}}$) with rate $\beta$. A proportion $h_{\mathrm{S}}$ $(0 \le h_{\mathrm{S}} \le 1)$) of newly infected individuals from $S_{\mathrm{H}}$ has severe disease, so $1-h_{\mathrm{S}}$ represents the proportion of becoming mildly symptomatic and infected from $S_{\mathrm{H}}$. Similarly, $l_{\mathrm{S}}$ represents the proportion of newly infected individuals derived from $S_{\mathrm{L}}$with severe disease. Since the $S_{\mathrm{L}}$ population has severity-reducing immunity, the proportion that acquires severe disease ($I_{\mathrm{S}}$) from $S_{\mathrm{L}}$ is less than that from $S_{\mathrm{H}}$ (i.e., $l_{\mathrm{S}} < h_{\mathrm{S}}$). This immunity wanes with rate $\omega_{S_{\mathrm{L}}\to S_{\mathrm{H}}}$. Infected individuals can recover with rate $\gamma$, so $1/\gamma$ represents the mean infectious period until recovery (or hospitalized). Recovered individuals have infection-preventing immunity, which wanes with rate $\omega_{R\to S_{\mathrm{L}}}$. $S_{\mathrm{H}}$ and $S_{\mathrm{L}}$ can also obtain infection-preventing immunity by vaccination with rate $\nu$.

#### The derivation of steady states

To investigate the number of each compartment in the model after stabilization, we explicitly derive the formulae for the steady-state values, which allows us to obtain the steady-state values without performing numerical simulations of the differential equations. Let $S_{\mathrm{T}} = S_{\mathrm{H}} + S_{\mathrm{L}}$, $I_{\mathrm{T}} = I_{\mathrm{S}} + I_{\mathrm{M}}$, and the $\infty$ at the subscript of a variable denotes the steady-state value of the variable. We can rewrite the differential equations using $S_{\mathrm{T}}$ and $I_{\mathrm{T}}$ as follows.

$$\frac{dS_{\mathrm{T}}}{dt} = -\beta\frac{S_{\mathrm{T}}I_{\mathrm{T}}}{N} - \nu S_{\mathrm{T}} + \omega_{R\to S_{\mathrm{L}}}R,$$
$$\frac{dI_{\mathrm{T}}}{dt} = \beta\frac{S_{\mathrm{T}}I_{\mathrm{T}}}{N} - \gamma I_{\mathrm{T}},$$
$$\frac{dR}{dt} = \gamma I_{\mathrm{T}} + \nu S_{\mathrm{T}} - \omega_{R\to S_{\mathrm{L}}}R.$$

The steady state of the model can be derived by setting $\frac{dS_{\mathrm{T}}}{dt} = \frac{dI_{\mathrm{T}}}{dt} = \frac{dR}{dt} = 0$. From these, we get

$$S_{\mathrm{T},\infty}\left(\beta\frac{I_{\mathrm{T},\infty}}{N} - \nu\right) = \omega_{R\to S_{\mathrm{L}}}R_{\infty}.$$
$$I_{\mathrm{T},\infty}\left(\beta S_{\mathrm{T},\infty} - \gamma N\right) = 0,$$
$$R_{\infty} = \frac{1}{\omega_{R\to S_{\mathrm{L}}}}(\gamma I_{\mathrm{T},\infty} + \nu S_{\mathrm{T},\infty})$$

From $I_{\mathrm{T},\infty}\left(\beta S_{\mathrm{T},\infty} - \gamma N\right) = 0$, $I_{\mathrm{T},\infty} = 0$ or $S_{\mathrm{T},\infty} = \frac{\gamma}{\beta}N$.

(i) If $I_{\mathrm{T},\infty} = 0$ then $R_\infty = \frac{\nu}{\omega_{R\to S_\mathrm{L}}} S_{\mathrm{T},\infty}$ and so $R_\infty = \frac{\nu}{\omega_{R\to S_\mathrm{L}}+\nu} N$ and $S_{\mathrm{T},\infty} = \frac{\omega_{R\to S_\mathrm{L}}}{\omega_{R\to S_\mathrm{L}}+\nu} N$ because $S_{\mathrm{T},\infty} + R_\infty = N$. This situation represents the extinction of the disease (i.e., disease-free equilibrium) as there is no infected individual in the system. From a simple stability analysis, it can be shown that this extinction occurs when $\frac{\beta}{\gamma} < 1 + \frac{\nu}{\omega_{R\to S_\mathrm{L}}}$, meaning that daily vaccination is high enough to counteract its waning rate and the transmissibility of the disease.

(ii) If $I_{\mathrm{T},\infty} \neq 0$, i.e., $\frac{\beta}{\gamma} \geq 1 + \frac{\nu}{\omega_{R\to S_\mathrm{L}}}$, $S_{\mathrm{T},\infty} = \frac{\gamma}{\beta} N$. From the conservation $I_{\mathrm{T},\infty} + S_{\mathrm{T},\infty} + R_\infty = N$, we have

$$I_{\mathrm{T},\infty} = \frac{1}{\beta(\omega_{R\to S_\mathrm{L}} + \gamma)} \left(\omega_{R\to S_\mathrm{L}}\beta - \omega_{R\to S_\mathrm{L}}\gamma - \nu\gamma\right) N,$$

$$R_\infty = N - S_{\mathrm{T},\infty} - I_{\mathrm{T},\infty}.$$

From the differential equation for the variable $S_\mathrm{L}$: $\frac{dS_\mathrm{L}}{dt} = -\beta \frac{(I_\mathrm{S}+I_\mathrm{M})}{N} S_\mathrm{L} - \nu S_\mathrm{L} - \omega_{S_\mathrm{L}\to S_\mathrm{H}} S_\mathrm{L} + \omega_{R\to S_\mathrm{L}} R$, $S_{\mathrm{L},\infty} = \frac{\omega_{R\to S_\mathrm{L}} R_\infty}{\beta \frac{I_{\mathrm{T},\infty}}{N} + \nu + \omega_{S_\mathrm{L}\to S_\mathrm{H}}}$ and $S_{\mathrm{H},\infty} = S_{\mathrm{T},\infty} - S_{\mathrm{L},\infty}$. Finally, from the differential equations for the variables $I_\mathrm{S}$ and $I_\mathrm{M}$, we can derive the steady states:

$$I_{\mathrm{S},\infty} = \frac{\beta}{\gamma} \frac{I_{\mathrm{T},\infty}}{N} (h_\mathrm{S} S_{\mathrm{H},\infty} + l_\mathrm{S} S_{\mathrm{L},\infty}) \text{ and } I_{\mathrm{M},\infty} = \frac{\beta}{\gamma} \frac{I_{\mathrm{T},\infty}}{N} ((1-h_\mathrm{S}) S_{\mathrm{H},\infty} + (1-l_\mathrm{S}) S_{\mathrm{L},\infty}).$$

**The derivation of a reproduction number incorporating vaccination ($R_\mathrm{v}$) and the basic reproduction number ($R_0$)**

In this section, we derive two reproduction numbers: one incorporating vaccination and the basic reproduction number[1]. The system has a disease-free equilibrium $x_0 = (S_\mathrm{H}^*, S_\mathrm{L}^*, 0, 0, R^*) = (S_\mathrm{H}^*, \frac{\nu}{\omega_{S_\mathrm{L}\to S_\mathrm{H}}} S_\mathrm{H}^*, 0, 0, \frac{\nu}{\omega_{R\to S_\mathrm{L}}+\nu} N)$ where $S_\mathrm{H}^* = \frac{\omega_{S_\mathrm{L}\to S_\mathrm{H}}}{\omega_{S_\mathrm{L}\to S_\mathrm{H}}+\nu} \frac{\omega_{R\to S_\mathrm{L}}}{\omega_{R\to S_\mathrm{L}}+\nu} N$. Let $x$ be the compartments, $(S_\mathrm{H}, S_\mathrm{L}, I_\mathrm{S}, I_\mathrm{M}, R)^\mathrm{T}$. We define the matrix of new infection, $\mathcal{F}(x)$, and the matrix of the transitions outward from the two infected compartments (i.e., $I_\mathrm{S}$ and $I_\mathrm{M}$), $\mathcal{V}(x)$, by

$$\mathcal{F}(x) = \begin{pmatrix} \mathcal{F}_1(x) \\ \mathcal{F}_2(x) \end{pmatrix} = \begin{pmatrix} \beta \frac{(I_\mathrm{S}+I_\mathrm{M})}{N} (h_\mathrm{S} S_\mathrm{H} + l_\mathrm{S} S_\mathrm{L}) \\ \beta \frac{(I_\mathrm{M}+I_\mathrm{M})}{N} ((1-h_\mathrm{S}) S_\mathrm{H} + (1-l_\mathrm{S}) S_\mathrm{L}) \end{pmatrix}, \mathcal{V}(x) = \begin{pmatrix} \mathcal{V}_1(x) \\ \mathcal{V}_2(x) \end{pmatrix} = \begin{pmatrix} \gamma I_\mathrm{S} \\ \gamma I_\mathrm{M} \end{pmatrix}.$$

The next generation matrix $K$ is defined as $FV^{-1}$ where $F = \left[\frac{\partial \mathcal{F}_i(x_0)}{\partial x_j}\right]$ and $V = \left[\frac{\partial \mathcal{V}_i(x_0)}{\partial x_j}\right]$ for $j = 3$ and $4$, which represent the infected compartments $I_\mathrm{S}$ and $I_\mathrm{M}$, respectively, that is,

$$\mathrm{F} = \begin{bmatrix} \frac{\beta(h_\mathrm{S} S_\mathrm{H}^* + l_\mathrm{S} S_\mathrm{L}^*)}{N} & \frac{\beta(h_\mathrm{S} S_\mathrm{H}^* + l_\mathrm{S} S_\mathrm{L}^*)}{N} \\ \frac{\beta((1-h_\mathrm{S}) S_\mathrm{H}^* + (1-l_\mathrm{S}) S_\mathrm{L}^*)}{N} & \frac{\beta((1-h_\mathrm{S}) S_\mathrm{H}^* + (1-l_\mathrm{S}) S_\mathrm{L}^*)}{N} \end{bmatrix} \text{ and } \mathrm{V} = \begin{bmatrix} \gamma & 0 \\ 0 & \gamma \end{bmatrix}.$$

Thus, the next generation matrix $K$ is calculated as

$$K = \mathrm{FV}^{-1} = \begin{bmatrix} \frac{\beta(h_\mathrm{S} S_\mathrm{H}^* + l_\mathrm{S} S_\mathrm{L}^*)}{\gamma N} & \frac{\beta(h_\mathrm{S} S_\mathrm{H}^* + l_\mathrm{S} S_\mathrm{L}^*)}{\gamma N} \\ \frac{\beta((1-h_\mathrm{S}) S_\mathrm{H}^* + (1-l_\mathrm{S}) S_\mathrm{L}^*)}{\gamma N} & \frac{\beta((1-h_\mathrm{S}) S_\mathrm{H}^* + (1-l_\mathrm{S}) S_\mathrm{L}^*)}{\gamma N} \end{bmatrix}.$$

Finally, the basic reproduction number $R_\mathrm{v}$ is given by the spectral radius (i.e., the maximum eigenvalue), $\rho(K)$, of the next generation matrix $K$[1]. Thus,

$$R_\mathrm{v} = \rho(K) = \frac{\beta(h_\mathrm{S} S_\mathrm{H}^* + l_\mathrm{S} S_\mathrm{L}^*) + \beta((1-h_\mathrm{S}) S_\mathrm{H}^* + (1-l_\mathrm{S}) S_\mathrm{L}^*)}{\gamma N} = \frac{\beta(S_\mathrm{H}^* + S_\mathrm{L}^*)}{\gamma N} = \frac{\omega_{R\to S_\mathrm{L}}}{\omega_{R\to S_\mathrm{L}}+\nu} \frac{\beta}{\gamma}.$$

To derive the basic reproduction number ($R_0$), we let $x_0$ be the state $(S_{\mathrm{H}}^*, S_{\mathrm{L}}^*, 0,0,0)$ at which the whole populations are susceptible. Then through the same derivation, $R_0 = \frac{\beta}{\gamma}$.

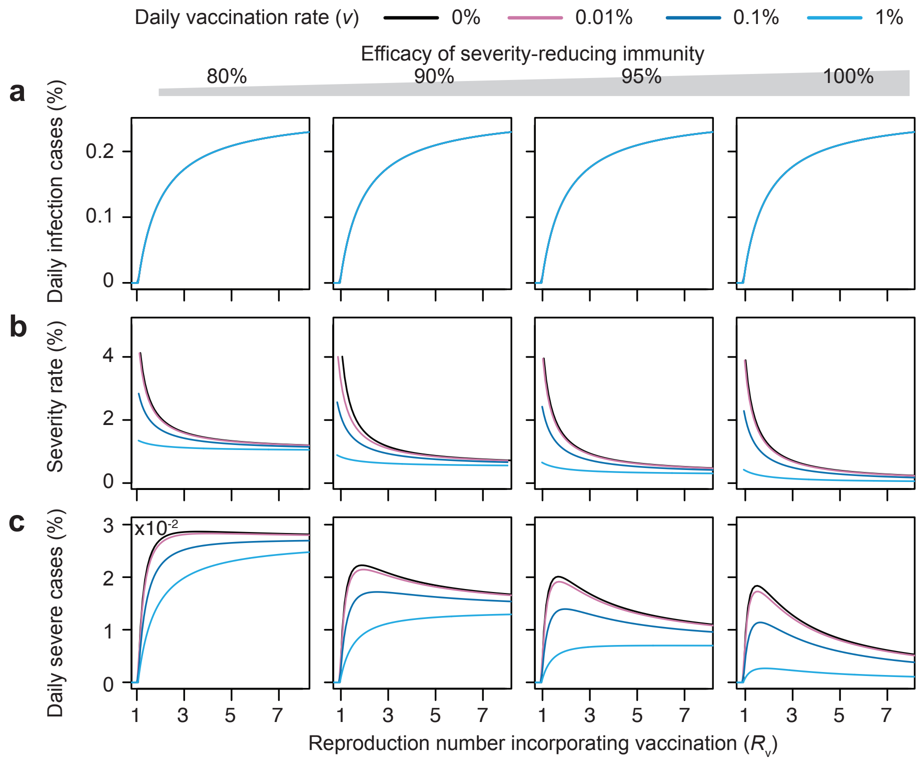

**Supplementary Fig. 1. Parallel figure of Fig. 2 generated with varying the reproduction number incorporating vaccination ($R_v$). a** The percentage of daily infections, **b** severity rates, and **c** the percentage of daily severe cases at the steady state depending on the reproduction number incorporating vaccination, $R_v = \frac{\omega_{R\to S_L}}{\omega_{R\to S_L}+\nu} R_0$. Although $R_v$ is used instead of $R_0$, the major patterns such as an initial increase followed by a decrease of daily severe cases are preserved compared with Fig. 2. See Supplementary Table 2 for the parameter values.

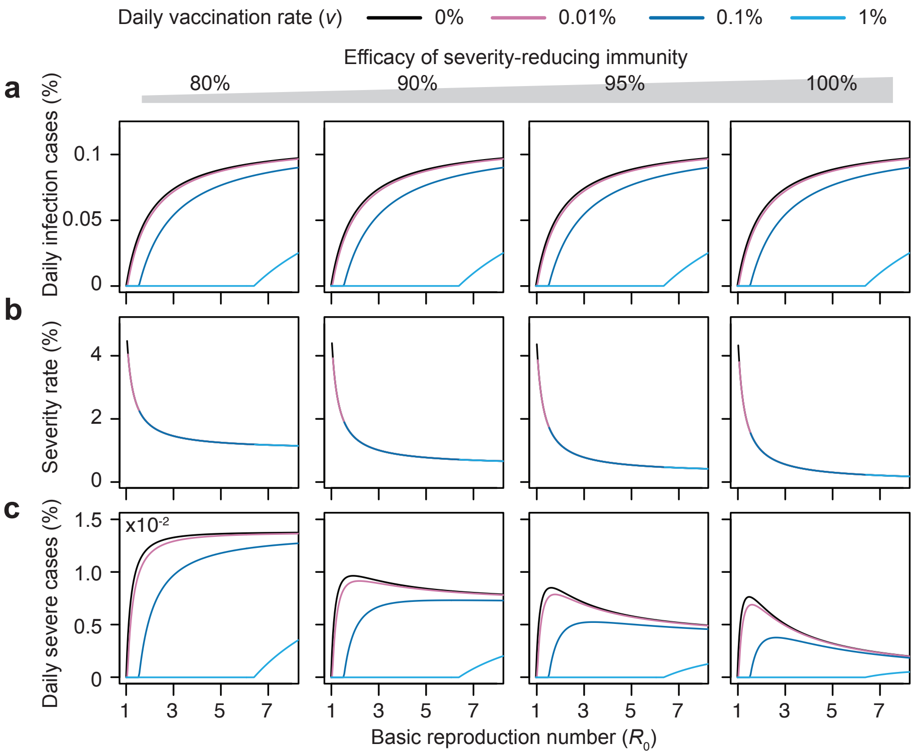

**Supplementary Fig. 2. Parallel figure of Fig. 2 generated by changing the recovery rate ($\gamma$) and immunity waning rates ($\omega_{R \to S_L}$ and $\omega_{S_L \to S_H}$). a** The percentage of daily infections, **b** severity rate, and **c** the percentage of daily severe cases at the steady state over the basic reproduction numbers ($\boldsymbol{R_0}$) with increased recovery rate ($\boldsymbol{\gamma}$) and reduced waning rates ($\boldsymbol{\omega_{R \to S_L}}$ and $\boldsymbol{\omega_{S_L \to S_H}}$). The major patterns, such as an initial increase followed by a decrease of the daily severe cases, are preserved compared with Fig. 2. See Supplementary Table 2 for the parameter values.

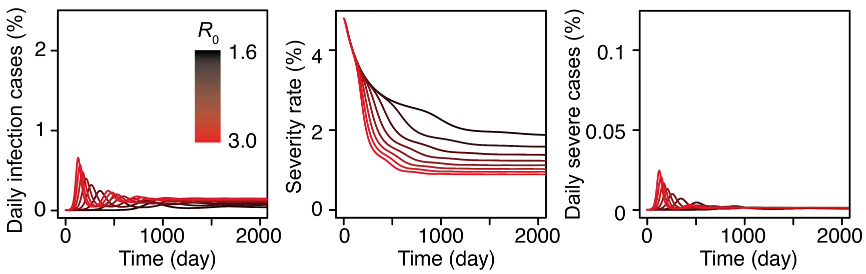

**Supplementary Fig. 3. The predicted dynamics of the proportion of daily cases among the whole population, the rate of severe disease among all infections, and the proportion of daily severe cases among the whole population, varying $R_0$ from 1.6 to 3.0 for initial population immunity of 50%.** See Supplementary Table 2 for the parameter values.

**Supplementary Table 1. Parameters of COVID-19 transmission model**

| Symbol | Description | Value | Reference |
|---|---|---|---|
| $\beta$ | Transmission rate | 0.1~2 | 2 |
| $h_{\mathrm{S}}$ (, $l_{\mathrm{S}}$) | Proportion with severe disease derived from $S_{\mathrm{H}}$ (, $S_{\mathrm{L}}$) to $I_{\mathrm{S}}$ | 0.05 (, 0~0.01) | 3-5 |
| $1/\gamma$ | Infectious period (days) | 4~10 days | 2,6 |
| $\nu$ | Immunization rate (1/days) | 0 ~ 0.01 | 7,8 |
| $\omega_{R\rightarrow S_{\mathrm{L}}}$ | Immune waning rate from $R$ to $S_{\mathrm{L}}$ (1/days) | 1/365 ~ 1/(365×3) | 8,9 |
| $\omega_{S_{\mathrm{L}}\rightarrow S_{\mathrm{H}}}$ | Immune waning rate from $S_{\mathrm{L}}$ to $S_{\mathrm{H}}$ (1/days) | 1/(365×3) ~ 1/(365×10) | 8,10,11 |

**Supplementary Table 2. The parameter values and initial conditions used in Figures**

| Parameters | Fig. 2 and Supplementary Fig. 1 | Fig. 3a-c & d-g & Supplementary Fig. 3 | Fig. 3h (orange, blue) | Supplementary Fig. 2 |
|---|---|---|---|---|
| $\beta$ | 0.1 ~ 0.8 | 0.16 ~ 0.3 | (0.3, 0.3) | 0.25 ~ 2 |
| $h_{\mathrm{S}}$ | 0.05 | 0.05 | $(5, 2)\times10^{-2}$ | 0.05 |
| $l_{\mathrm{S}}$ | 0 ~ 0.01 | 0.0025 | $(2.5, 1)\times10^{-3}$ | 0 ~ 0.01 |
| $\gamma$ | 0.1 | 0.1 | (0.1, 0.17) | 0.25 |
| $\nu$ | 0 ~ 0.01 | 0.001 | $(1, 1)\times10^{-3}$ | 0 ~ 0.01 |
| $\omega_{R\to S_{\mathrm{L}}}$ | 1/365 | 1/365 | (1/3, 1)×1/365 | 1/(365×1.5) |
| $\omega_{S_{\mathrm{L}}\to S_{\mathrm{H}}}$ | 1/(365×3) | 1/(365×3) | (1/10, 1/3)×1/365 | 1/(365×6) |
| Initial conditions $(S_{\mathrm{H}}, S_{\mathrm{L}}, I_{\mathrm{S}}, I_{\mathrm{M}}, R)$ | - | $(0.5\times10^{7}, 0, 250, 4750, 4.5\times10^{7})$ & $(4.0\times10^{7}, 0, 250, 4750, 1.0\times10^{7})$ & $(2.5\times10^{7}, 0, 250, 4750, 2.5\times10^{7})$ | $(4.0\times10^{7}, 0, 250, 4750, 1.0\times10^{7})$ | - |

### References for Supplementary materials

1 van den Driessche, P. Reproduction numbers of infectious disease models. *Infect. Dis. Model* **2**, 288-303, doi:10.1016/j.idm.2017.06.002 (2017).

2 Hellewell, J. *et al.* Feasibility of controlling COVID-19 outbreaks by isolation of cases and contacts. *Lancet Glob. Health* **8**, e488-e496, doi:10.1016/S2214-109X(20)30074-7 (2020).

3 Thomas, S. J. *et al.* Safety and Efficacy of the BNT162b2 mRNA Covid-19 Vaccine through 6 Months. *N. Engl. J. Med.* **385**, 1761-1773, doi:10.1056/NEJMoa2110345 (2021).

4 Abu-Raddad, L. J., Chemaitelly, H., Bertollini, R. & National Study Group for, C.-E. Severity of SARS-CoV-2 Reinfections as Compared with Primary Infections. *N. Engl. J. Med.* **385**, 2487-2489, doi:10.1056/NEJMc2108120 (2021).

5 Falsey, A. R. *et al.* Phase 3 Safety and Efficacy of AZD1222 (ChAdOx1 nCoV-19) Covid-19 Vaccine. *N. Engl. J. Med.* **385**, 2348-2360, doi:10.1056/NEJMoa2105290 (2021).

6 Lin, S.-N. *et al.* Effectiveness of potential antiviral treatments in COVID-19 transmission control: a modelling study. *Infect. Dis. Poverty* **10**, 53, doi:10.1186/s40249-021-00835-2 (2021).

7 Ritchie, H. *et al.* Coronavirus Pandemic (COVID-19). (2020). https://ourworldindata.org/coronavirus (accessed Jan 18, 2022)

8 Bubar, K. M. *et al.* Model-informed COVID-19 vaccine prioritization strategies by age and serostatus. *Science* **371**, 916-921, doi:10.1126/science.abe6959 (2021).

9 Townsend, J. P. *et al.* The durability of immunity against reinfection by SARS-CoV-2: a comparative evolutionary study. *Lancet Microbe* **2**, e666-e675, doi:10.1016/S2666-5247(21)00219-6 (2021).

10 Le Bert, N. *et al.* SARS-CoV-2-specific T cell immunity in cases of COVID-19 and SARS, and uninfected controls. *Nature* **584**, 457-462, doi:10.1038/s41586-020-2550-z (2020).

11 Yang, L. T. *et al.* Long-lived effector/central memory T-cell responses to severe acute respiratory syndrome coronavirus (SARS-CoV) S antigen in recovered SARS patients. *Clin. Immunol.* **120**, 171-178, doi:10.1016/j.clim.2006.05.002 (2006).
